# Plasma Proteomic Signatures of Physical Activity Provide Insights into Biological Impacts of Physical Activity and its Protective Role Against Dementia

**DOI:** 10.1101/2025.01.16.25320290

**Authors:** Gayatri Arani, Amit Arora, Shuai Yang, Jingyue Wu, Jennifer N. Kraszewski, Amy Martins, Alexandra Miller, Zebunnesa Zeba, Ayan Jafri, Chengcheng Hu, Leslie V. Farland, Jennifer W. Bea, Dawn K. Coletta, Daniel H. Aslan, M Katherine Sayre, Pradyumna K Bharadwaj, Madeline Ally, Silvio Maltagliati, Mark H C Lai, Rand Wilcox, Eco de Geus, Gene E. Alexander, David A. Raichlen, Yann C. Klimentidis

**Affiliations:** Department of Epidemiology and Biostatistics, College of Public Health, University of Arizona, Tucson, AZ, USA; Department of Biomedical Informatics, College of Health Solutions, Arizona State University, Tempe, AZ, USA; College of Medicine, University of Arizona, Tucson, AZ, USA; Division of Epidemiology, Biostatistics, and Environmental Health, School of Public Health, University of Memphis, Memphis, TN, USA; Department of Health Promotion Sciences, University of Arizona, Tucson, AZ, USA; University of Arizona Cancer Center, Tucson, AZ, USA; Department of Physiology, University of Arizona, Tucson, AZ, USA; Department of Medicine, Division of Endocrinology, University of Arizona, Tucson, AZ, USA; Department of Clinical and Translational Genomics, University of Arizona, Tucson, AZ, USA; Center for Disparities in Diabetes, Obesity and Metabolism, University of Arizona, Tucson, AZ, USA; Department of Anthropology, University of Southern California, Los Angeles, CA, USA; Department of Anthropology, University of California Santa Barbara, Santa Barbara, CA, USA; Department of Psychology, University of Arizona, Tucson, AZ, USA; University of Grenoble Alpes, SENS, Grenoble 38000, France; Department of Psychology, University of Southern California, Los Angeles, CA, United States; Department of Biological Psychology, Vrije Universiteit, Amsterdam, the Netherlands; Amsterdam Public Health Research Institute, Amsterdam UMC, Amsterdam, the Netherlands; BIO5 Institute, University of Arizona, Tucson, AZ, USA; Evelyn F. McKnight Brain Institute, University of Arizona, Tucson, AZ, USA; Department of Psychiatry, University of Arizona, Tucson, AZ, USA; Neuroscience Graduate Interdisciplinary Program, University of Arizona, Tucson, USA; Arizona Alzheimer’s Consortium, Phoenix, AZ, USA; Human and Evolutionary Biology Section, Department of Biological Sciences, University of Southern California, Los Angeles, CA, USA; Genetics Graduate Interdisciplinary Program, University of Arizona, Tucson, AZ, USA

## Abstract

Physical activity (PA), including sedentary behavior, is associated with many diseases, including Alzheimer’s disease and all-cause dementia. However, the specific biological mechanisms through which PA protects against disease are not entirely understood. To address this knowledge gap, we first assessed the conventional observational associations of three self-reported and three device-based PA measures with circulating levels of 2,911 plasma proteins measured in the UK Biobank (n_max_=39,160) and assessed functional enrichment of identified proteins. We then used bi-directional Mendelian randomization (MR) to further evaluate the evidence for causal relationships of PA with protein levels. Finally, we performed mediation analyses to identify proteins that may mediate the relationship of PA with incident all-cause dementia. Our findings revealed 41 proteins consistently associated with all PA measures and 1,027 proteins associated with at least one PA measure. Both conventional observational and MR study designs converged on proteins that appear to increase as a result of PA, including integrin proteins such as ITGAV and ITGAM, as well as MXRA8, CLEC4A, CLEC4M, GFRA1, and ADGRG2; and on proteins that appear to decrease as a result of PA such as LEP, LPL, INHBC, CLMP, PTGDS, ADM, OGN, and PI3. Functional enrichment analyses revealed several relevant processes, including cell-matrix adhesion, integrin-mediated signaling, and collagen binding. Finally, several proteins, including GDF15, ITGAV, HPGDS, BCAN, and MENT, were found to mediate the relationship of PA with all-cause dementia, implicating processes such as synaptic plasticity, neurogenesis and inflammation, through which PA protects against dementia. Our results provide insights into how PA may affect biological processes and protect from all-cause dementia, and provide avenues for future research into the health-promoting effects of PA.

## INTRODUCTION

Physical activity (PA) and sedentary behavior (SB), hereafter collectively referred to as PA, are associated with health outcomes such as cardiovascular function, weight management, cognitive function, and mental health.^1^ There are well-established associations between PA and risk of several chronic diseases such as heart disease, type-2 diabetes, depression, anxiety, osteoporosis, and cancer.^2,3^ PA is also increasingly recognized for its potential role in the risk and progression of dementia.^4–8^

Regular PA may enhance neuroplasticity and cognitive functioning through improved cerebral blood circulation, stimulation of neural tissue development, and the release of other immunological and endocrine factors.^9–11^ Additionally, PA improves metabolic functioning, in part, by reducing systemic inflammation and oxidative stress, which are pathological mechanisms also implicated in all-cause dementia.^12–15^ Although the neurocognitive benefits of PA are often observed, the precise mechanisms and biological underpinnings of this protection are not well understood.^12,16^ Investigating the biological pathways that underlie the association between PA and all-cause dementia can increase opportunities for the precise and effective prevention and management of dementia.^12^

Investigating circulating plasma protein levels offers a promising avenue to uncover the biological mechanisms underlying the health-promoting effects of PA.^16,17^ These insights could pave the way for innovative applications in clinical practice, enabling more targeted and effective interventions.^16,17^ However, previous human studies examining the relationship between PA and plasma protein levels have been based on relatively small sample sizes or small sets of measured proteins.^18–22^ For example, Stattin et al.^18^ examined 184 proteins among 6,500 individuals. Moreover, only self-reported PA measures are typically examined.^18,19^ While substantial progress has been made in understanding the proteomic changes associated with cardiovascular health and PA, there remains a critical gap in linking these molecular changes to cognitive outcomes such as dementia.^17–24^ For instance, a study by Robbins et al.^25^ demonstrated how plasma proteomic changes in response to exercise training are associated with cardiorespiratory fitness (CRF) adaptations, highlighting the diverse physiological pathways through which PA can confer cardiorespiratory benefits. Furthermore, while previous research has attempted to find associations between PA and plasma protein levels, conventional observational studies may be biased by unmeasured and residual confounding factors, and reverse causation.^26^ As a result, alternative study designs such as Mendelian randomization (MR) can help to assess causal relationships.^26–28^

We performed the most extensive study to date testing the associations of PA with the levels of 2,911 plasma proteins, using self-reported and device-based measures (n_max=_39,160) from the UK Biobank. We compared the evidence for causality across the different types of PA measures and two study designs: a conventional observational study and a bi-directional MR study. We then assessed the functional enrichment of identified proteins and assessed whether any specific proteins mediate the relationship between PA and all-cause dementia in the UK Biobank (Figure 1).

**Figure 1:**
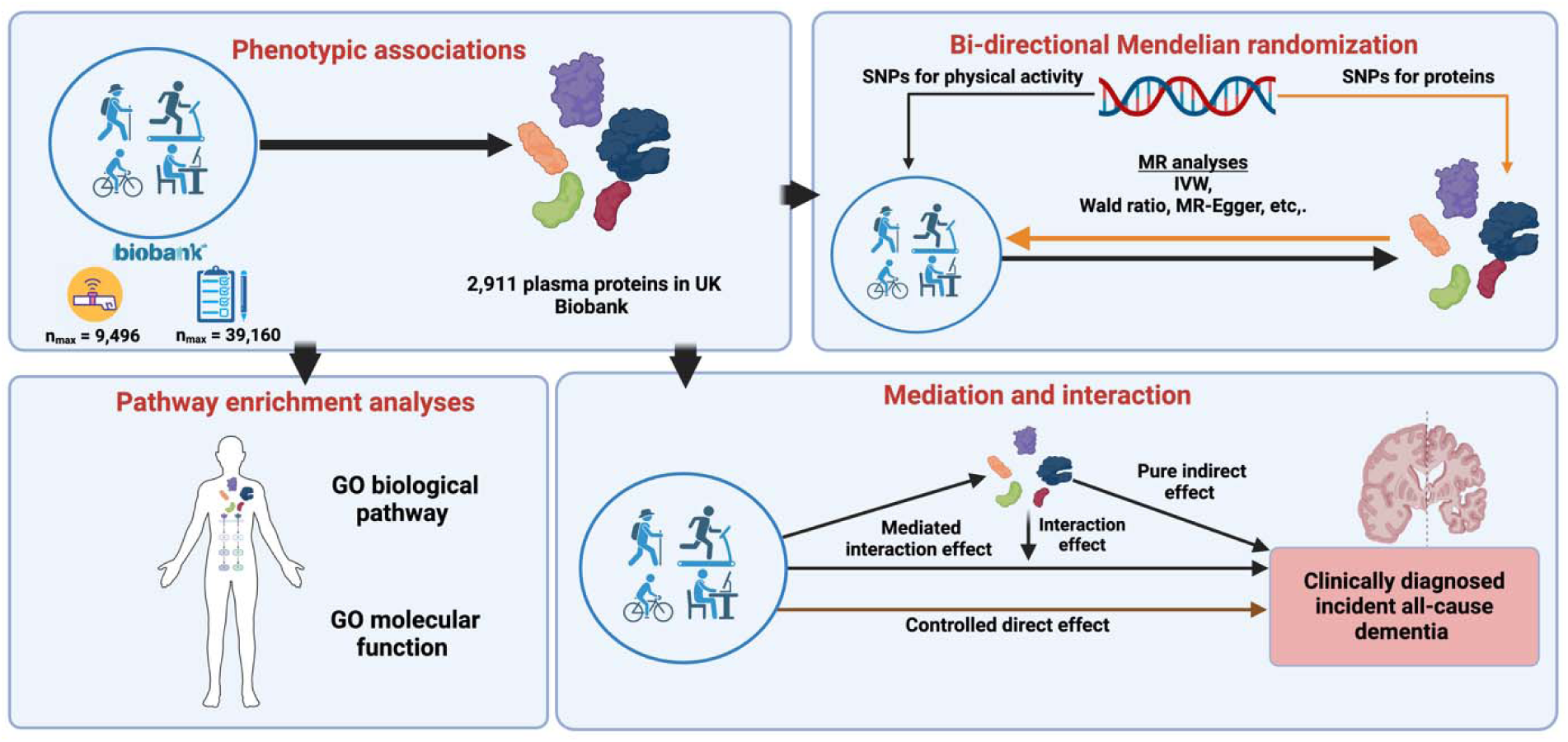
Study design and analytical framework for investigating the relationship of physical activity and sedentary behaviors with plasma proteins and incident all-cause dementia. The framework consists of four main components: (1) Phenotypic (conventional observational) associations: Analysis of associations between PA measures and 2,911 plasma proteins from the UK Biobank, with sample sizes ranging from n = 9,496 to n = 39,160. (2) Pathway enrichment analyses: Identification of protein signaling pathways, Gene Ontology (GO) biological processes, and GO molecular functions enriched among the associated proteins. (3) Bi-directional Mendelian randomization: Examination of causal relationships between PA and proteins using genetic instruments (SNPs) for both PA and protein levels, employing Mendelian Randomization (MR) methods such as Inverse Variance Weighted (IVW), Wald Ratio, and MR-Egger. (4) Mediation and interaction analyses: Assessment of the direct, indirect, mediated interaction, and interaction effects of PA-associated proteins on the risk of clinically diagnosed incident all-cause dementia. (Created with <u>BioRender.com</u>)

## METHODS

### UK Biobank proteomics

The UK Biobank is a large prospective study with over ∼ 500,000 participants aged 39-73 years at the time of recruitment between 2006 and 2010.^29^ Between November 2020 and March 2021, the Pharma Proteomics Project (PPP) consortium selected 54,219 representative participants through age, sex, and study center stratification for the proteomics sub-study.^30^ Blood samples were obtained at each participant’s baseline visit. The time of day of blood sample collection varied for each individual.^30^ UK Biobank has approval from the North West Multi-centre Research Ethics Committee (MREC) as a Research Tissue Bank (RTB).

Between May 2021 -November 2022, plasma profiling was performed using Olink Explore 3072 (Uppsala, Sweden) technology, in which a matched pair of antibodies labeled with unique complementary oligonucleotides (proximity probes) bind to the respective target proteins in each sample.^30^ The oligonucleotides come into proximity and hybridize with each other to enable DNA amplification of the protein signal, which is quantified on a next-generation sequencing read-out, as detailed elsewhere.^30^ Antibodies for 2,923 unique plasma proteins were available in the UK Biobank PPP.^30^ The protein panel covers major biological pathways in inflammation, oncology, cardiometabolic, and neurological domains.^30^ The performance of each protein assay is validated for specificity, sensitivity, dynamic range, precision, scalability, detectability, and endogenous interference.^30^ Additional details on sample batching, data pre-processing, and quality checking were previously published.^29^ Quality control of the proteomic data for the current study was performed in R (version 4.33)^31^ by excluding proteins with more than 20% missing values (n=12) and samples with more than 50% missing values, leaving a set of 2,911 proteins for analyses.

### Conventional observational associations of PA with protein levels

We used linear regression to examine the cross-sectional associations of six PA-related variables with each plasma protein. Three PA variables were self-reported: Moderate-to-Vigorous Physical Activity (MVPA), Vigorous Physical Activity (VPA), and Strenuous Sports and Other Exercises (SSOE). As previously described, these were derived from questionnaire responses obtained during the UK Biobank baseline assessment (2006-2010).^32^ Additionally, three device-based PA measures were assessed in 2014 for associations with plasma protein levels measured 4-8 years earlier at the baseline visit.^32^ A subset of 103,684 participants agreed to wear a 3-axis logging accelerometer (AX3; Activity) for 24 hours per day for seven days on their dominant wrist, from 2013 – 2015. These device-based measures were: average acceleration (AvgAcc), the fraction of accelerations >425 milligravities (FracAcc>425), and sedentary behavior (AccSed) derived from a machine-learning model^33^ as previously applied.^7^ The questions for self-reported PA measurements, participant device-wear information, and variable construction were created and transformed as previously described.^32^ We applied consistent exclusion criteria and data quality control measures to the accelerometry-based measures. Individuals with at least 3 days of data were included in the study.^7,32^

The numbers of participants with proteomic data and self-reported PA measures were as follows: MVPA: n=40,787, VPA n=10,502 physically active and 18,092 controls, SSOE: n=13,179 physically active and 25,770 controls) and device-based measures (AvgAcc: n=9,851, FracAcc>425: n= 9,792, AccSed: n=9,806). We conducted linear regression for the independent relationship between each PA variable as the exposure and each protein as the outcome, resulting in a total of 6 × 2,911 regressions. All proteins, along with MVPA and AvgAcc, were inverse normalized before analysis to ensure normality and comparability across variables. Each model assessing the PA-protein association was adjusted for age (at baseline or device use), sex, season (questionnaire or device use), race and ethnicity, body fat percentage (measured by a Tanita BC418MA bioimpedance scale), alcohol intake (drinks per week), smoking status (never, former, current), education (college or higher vs. less), and the presence of major health conditions (cancer, type-2 diabetes, hypertension). All statistical analyses of conventional observational associations were performed using R software (version 4.3.3). We selected all PA-protein pairs with a p < 1.7 × 10^-5^ as statistically significant, based on a Bonferroni correction for 2,911 tests. Although 6 PA measures were tested, we consider this a conservative correction given the substantial correlations among proteins and among PA measures.

### Pathway and functional enrichment analyses

We conducted gene enrichment analyses for the gene set corresponding to the proteins significantly associated with each PA measure and the set of proteins significantly associated with all six PA measures. We analyzed DAVID gene enrichment and functional annotation using the complete set of 2,911 Olink 3072 proteins as the background.^34–35,36^ We derived enrichment terms for Gene Ontology (GO) biological process, and GO molecular function. A Benjamini-Hochberg correction was used to assess whether our identified genes are significantly enriched for specific functional pathways.

### Bi-directional Mendelian randomization

We performed bi-directional MR for PA and protein levels for all pairs with a significant conventional observational association. We assessed PA exposure SNPs selected at the p < 5 × 10^-8^ and p < 5 × 10^-6^ thresholds. The stricter threshold (p < 5 × 10^-8^) ensured the inclusion of high-confidence genetic variants, while the slightly relaxed threshold (p < 5 × 10^-6^) allowed for additional variants since some PA exposures had fewer than 5 SNPs at the p < 5 × 10^-8^ threshold. We used GWAS of the same set of PA variables described above^32^, plus two additional self-reported measures based on larger sample sizes^37^: leisure screen time (n= 526,725) and MVPA during leisure time (n= 608,595). We used publicly available GWAS summary statistics reported in Sun et al. ^38^ to genetically-instrument protein levels. Specifically, we used the GWAS based on the European ancestry subset in the UK Biobank PPP (n= 34,557) since this corresponds to the ancestry used in the PA GWAS. Additionally, we performed MR with each protein as the exposure, limiting our exposure SNPs to cis-pQTLs within one Mb up and downstream of the transcription start site for the respective protein-coding gene to reduce the potential for pleiotropic variants outside the protein-encoding gene.^39^ All MR statistical analyses were performed in R (version 4.2) using the ‘TwoSampleMR’ package.^40^ Clumping for MR was performed with Plink 1.9 using the ‘EUR’ subpopulation from 1,000 Genomes as a reference. We first assessed results from the inverse variance weighted (IVW) method. When only one SNP exposure instrument was available, we applied the Wald ratio method and reported those results. To strengthen our findings, we examined the results of sensitivity analyses, including MR-Egger regression to account for directional pleiotropy, the weighted median method to account for the presence of invalid SNPs, and a mode-based estimator that grouped similar SNPs to minimize the impact of outliers. F-statistics for PA exposure instruments are all >10, as presented elsewhere.^41,42^ We selected all PA-protein pairs that showed evidence from MR for a causal effect in either direction, using a pre-set significance criterion of p < 4.9 × 10^-5^. This criterion was chosen based on a Bonferroni correction for the number of proteins tested. The significant protein-PA pairs were forwarded to the colocalization analysis.

### Colocalization

We performed colocalization analyses for PA-protein pairs that showed significant associations in MR analyses to rule out the potential of pleiotropy that could bias the MR results. We used the ‘*coloc’* package in R.^43^ We applied the ‘coloc.abf’ function using a genomic region of +/- 250kb around the variants to assess the colocalization of PA measures and proteins.^44^ We used the default threshold for colocalization evidence using the posterior probability (PP) of hypothesis 4 (H_4:_ PA and protein have one shared common associated/causal variant for both measures). A PP of H4 > 0.8 was considered as nominal evidence of colocalization.

### Mediation and interaction analyses (4-way decomposition)

Plasma proteins significantly associated with at least one PA measure (n=1,027) were analyzed using a four-way decomposition method to estimate the extent to which specific protein(s) mediate the PA-dementia relationship. Only participants aged 55 or older without a prior dementia diagnosis at the baseline visit were included in these analyses. Two primary models were utilized in the four-way decomposition analysis. For the exposure-mediator relationship, a linear regression model was used to estimate the association between PA (independent variable) and each selected protein mediator (dependent variable). This model was adjusted for the following covariates: age, sex, season, race and ethnicity, education, alcohol consumption, smoking status, presence of chronic diseases, and body fat percentage. A Cox proportional hazards regression model was employed to examine the association between each protein mediator and the incidence of all-cause dementia. Follow-up time and dementia incidence data were extracted from the UK Biobank. All-cause dementia was identified through diagnoses recorded in inpatient hospital records and death registry data, as previously detailed.^7^ Our definition encompassed all forms of dementia, including Alzheimer’s disease, vascular dementia, and other types. The follow-up period for dementia incidence began at the baseline assessment of self-reported PA (2006-2010) or at the time of device wear (2013-2015), depending on the specific analysis. Follow-up continued until the earliest occurrence of a dementia diagnosis, the last available health record, or the end of the study period in 2021, whichever occurred first. The four-way approach decomposes the total effect between PA and dementia into four main parameters: (1) Controlled Direct Effect (CDE): This parameter estimates the direct effect of a PA measure on the incidence of dementia, independent of the protein mediator.^45^ The protein is set to a fixed value for the entire study population to mimic the absence of the mediator. In this study, the fixed value of each protein was set to the mean of that protein. (2) Reference Interaction (Intref): This parameter indicates the effect of PA on the incidence of dementia due to only an interactive effect between the PA and the protein on dementia. (3) Mediation interaction (Intmed): This parameter refers to the effect of a PA measure on the incidence of dementia from both mediation by the protein and the interaction between PA and the protein. (4) Pure Indirect Effect (PIE): Also referred to as “mediated main effect,” this parameter estimates how much of the relationship between PA and dementia is explained by the influence of a given protein, which in turn influences dementia.^45^ Proteins with mediation (PIE) p < 4.9 × 10^-5^ were considered proteome-wide statistically significant. We also report the proportion of the total effect that consists of pure indirect effect. We used the *med4way* command in Stata 17 (StataCorp, College Station, TX) to perform the four-way decomposition analysis.^46^

## RESULTS

Our comprehensive findings are available through an interactive web platform ( https://github.com/klimentidis-lab/ProteomicsofPhysicalActivity2024.git), where it is also possible to visually browse the complete set of results with interactive volcano plots.

### Conventional observational associations of PA with proteins

Participants were 54% female and had a mean age of 56.8 years (Supplementary Table S1). Across all three self-reported (n_max_= 39,160) and three device-based (n_max_=9,496) PA measures (Supplementary Table S2), there were a total of 1,027 proteins (Supplementary Table S3) that were significantly associated with at least one measure, and 41 proteins (Supplementary Table S4) that were significantly associated with all six measures (Supplementary Tables S5-S10). Some of the strongest and most consistently associated proteins across self-report and device measures included increased levels of ITGAV, ITGAM, ITGA11, MXRA8, CA14, CLEC4A, MEGF10, MYOM3, MYL3, ADGRG2, ADAMTS8, and PON3, and decreased levels of LEP, GDF15, and IL6, among others (Figure 2).

**Figure 2:**
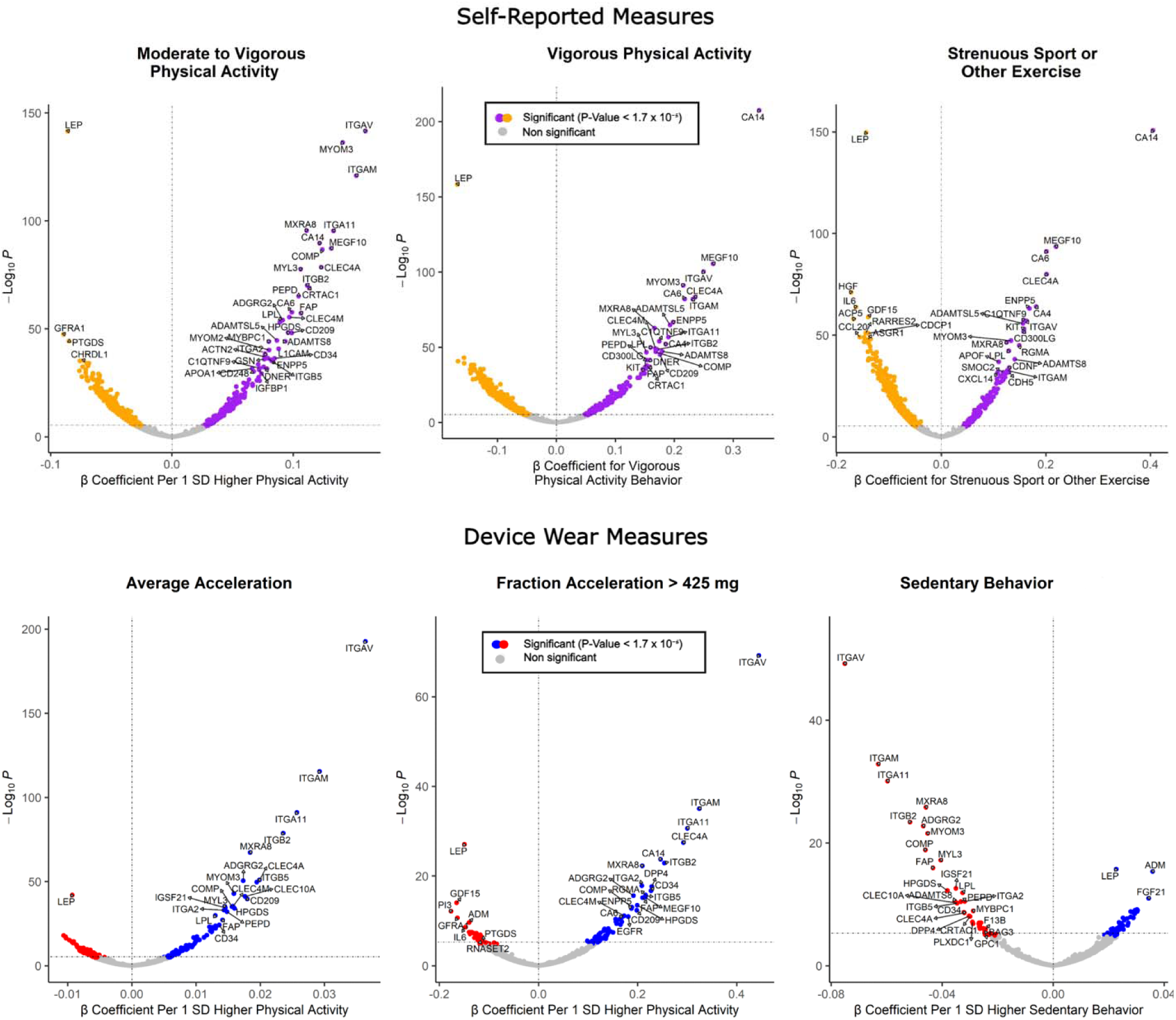
Volcano plots showing the associations of physical activity (PA) and sedentary behavior (SB) measures with protein levels. The six panels illustrate different PA measures, including self-reported measures (Moderate to Vigorous Physical Activity [MVPA], Vigorous Physical Activity [VPA], Strenuous Sport or Other Exercise [SSOE]) on the top row, and device-wear measures (Average Acceleration, Fraction Acceleration milligravities, Sedentary Behavior) on the bottom row. The y-axis in all panels represents the *-log10*(p) of the associations. For continuous PA measures, the x-axis shows the β coefficient per 1 standard deviation (SD) increase in PA. In contrast, for binary measures (VPA and SSOE), it represents the β coefficient per unit increase. Each point corresponds to a protein, with significant positive associations (p < 1.7 × 10^-5^, denoted by the gray horizontal dashed line) highlighted in purple (self-reported measures) and blue (device-wear measures). Significant negative associations are depicted in orange (self-reported measures) and red (device-wear measures).

### Pathway enrichment

The set of genes coding for the 41 proteins we found to be associated with every PA measure was enriched for several biological processes and molecular functions, including integrin-mediated signaling, cell-matrix adhesion, and collagen binding (Table 1). Carbohydrate binding and serine-type endopeptidase activity were significantly enriched in protein-coding genes specific to MVPA and VPA and to the set of 1,027 proteins associated with at least one PA measure (Supplementary Tables S11 & S12). Cell adhesion processes emerged as most prominently enriched for VPA, SSOE, and all device-based measures (Supplementary Table S11). The pathway enrichment analysis also revealed that cell adhesion mediated by integrins was consistently observed across four measures, including self-reported and device-based measures (Supplementary Table S11). Furthermore, proteins identified for AvgAcc and AccSed were found to participate in integrin-mediated signaling pathways (Supplementary Table S11). Integrin binding was recognized as a common pathway across the various device-based measures examined (Supplementary Table S11).

**Table 1:**
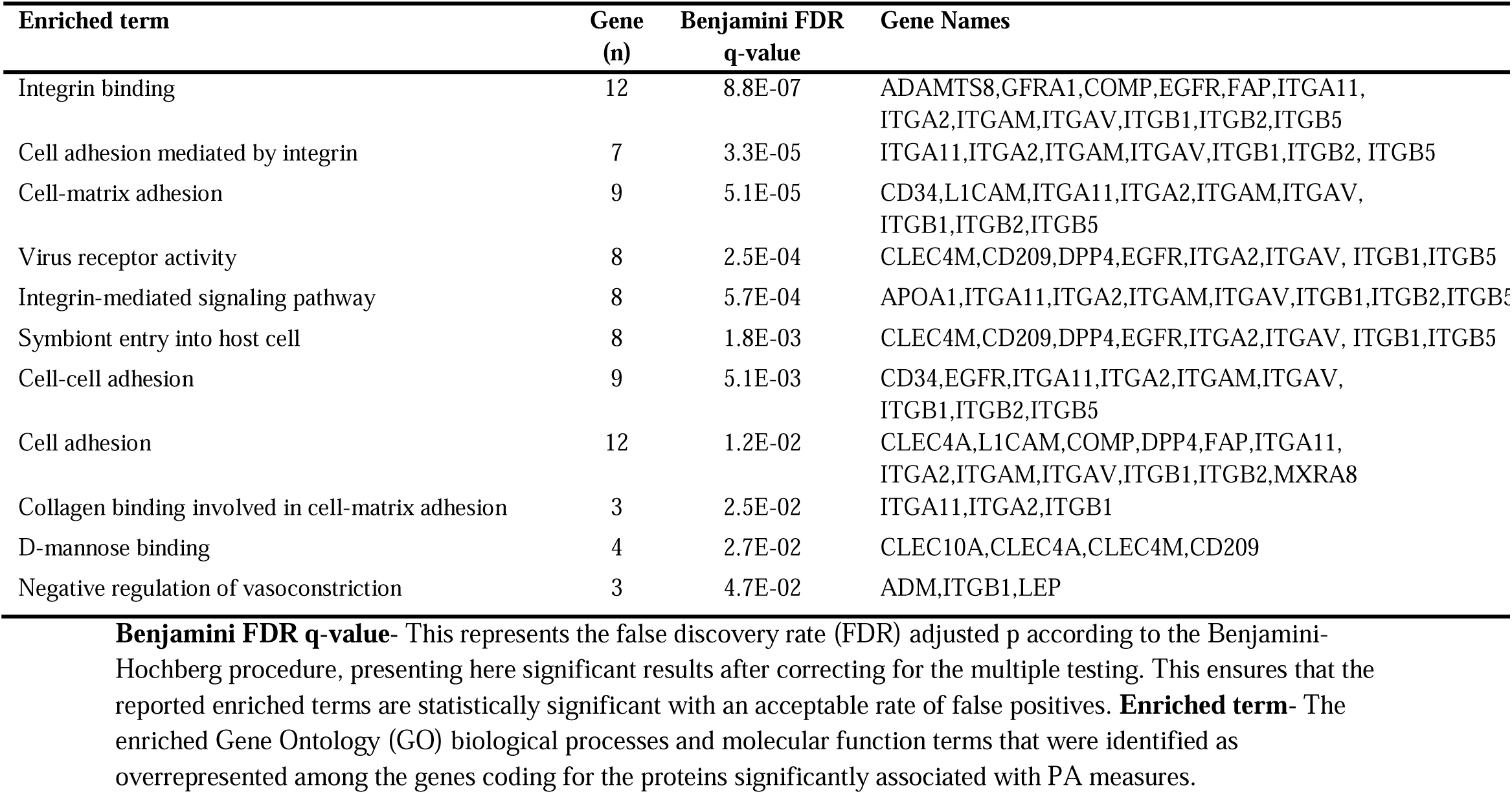
Enrichment of Gene Ontology biological process and molecular function terms (with Benjamini FDR <0.05) among the genes coding for the set of 41 proteins that were associated with all PA measures.

### Mendelian randomization

Of 1,027 proteins tested based on showing at least one significant (p < 1.7 × 10^−5^) association in the above conventional observational analyses, we found 194 significant (p < 4.9 × 10^-5^) MR IVW associations between PA exposure and protein level (with PA exposure SNPs at p < 5 × 10^-8^) and 280 proteins (IL6, LEP, ITGAV, MXRA8 and TNF were significantly associated with four or more measures) with the PA exposure SNPs at the p < 5 × 10^-6^ threshold (Figure 3). Three proteins (DUSP13, FGR, and RAB6A) and one protein (FGR) were significant in the reverse direction at the p < 5 × 10^-8^ and p < 5 × 10^-6^ thresholds for selecting the protein exposure SNPs, respectively. Using only cis-pQTL SNPs as protein exposure instruments revealed four (SLC9A3R2, DUSP13, GOLM2, CCL11) and three (SLC9A3R2, AP3B1, CCL11) proteins at the p < 5 × 10^-8^ and p < 5 × 10^-6^ thresholds, respectively. MR sensitivity analyses generally revealed directionally consistent estimates for most IVW top hits (Supplementary Tables S13 to S22). Based on our colocalization analyses, 110 distinct proteins and PA-related SNP pairs were observed to have a posterior probability of > 0.80 for a potential pleiotropic relationship (see Supplementary Table S23).

**Figure 3:**
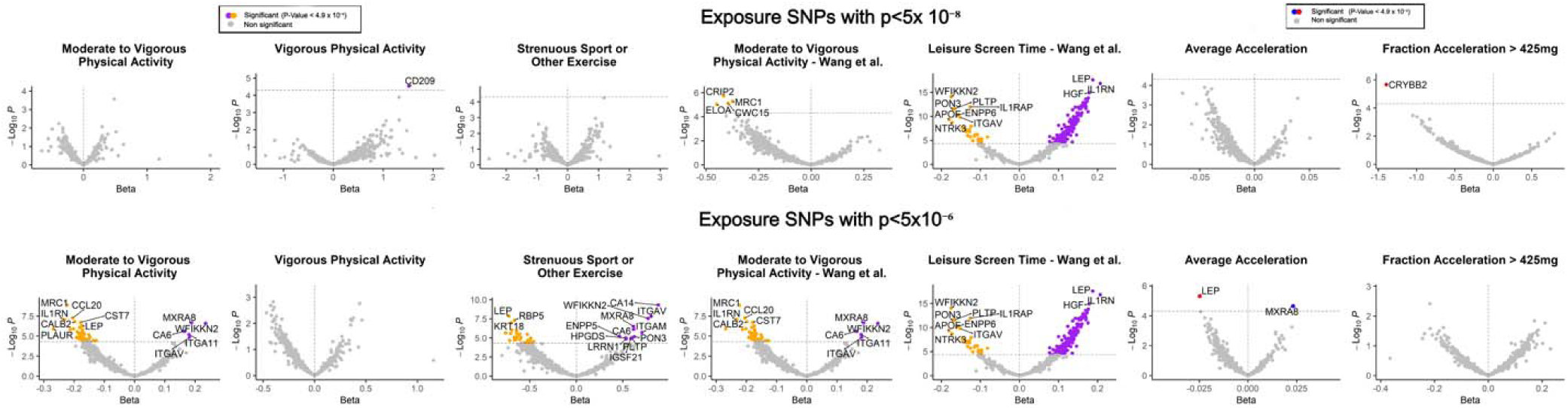
Volcano plots depicting Mendelian randomization results of genetically-predicted physical activity (PA) and sedentary behavior (SB) measures on protein levels, using IVW or Wald ratio methods. The top row shows results using exposure SNPs with p-values below the stringent genome-wide significance threshold (p < 5 × 10^-8^), while the bottom row displays results using exposure SNPs with associations at a more liberal genome-wide threshold (p < 5 × 10^-6^). Both rows highlight each PA and SB measure related to protein levels, with a statistical significance threshold of p < 4.9 × 10^-5^, indicated by the horizontal gray dashed line. Proteins with significant positive associations are shown in purple for self-reported measures and blue for device-wear measures, while proteins with significant negative associations are shown in orange and red.

To assess the consistency of the effect estimates of PA on protein levels across conventional observational and MR analyses, we plotted the beta coefficient estimates from the conventional observational analyses against the beta coefficient estimates from the MR analyses for each of the 1,027 proteins (Figure 4). When evaluating our results across the two study designs, we emphasized proteins exhibiting directionally consistent effects and statistically significant at p < 1 × 10^-3^ in both conventional observational and MR analyses. We used a more liberal threshold for statistical significance, considering the lower null likelihood of directionally consistent results in both designs. We identified ITGAV, MXRA8, and LEP as exhibiting consistent associations across study designs and PA measures.

**Figure 4.**
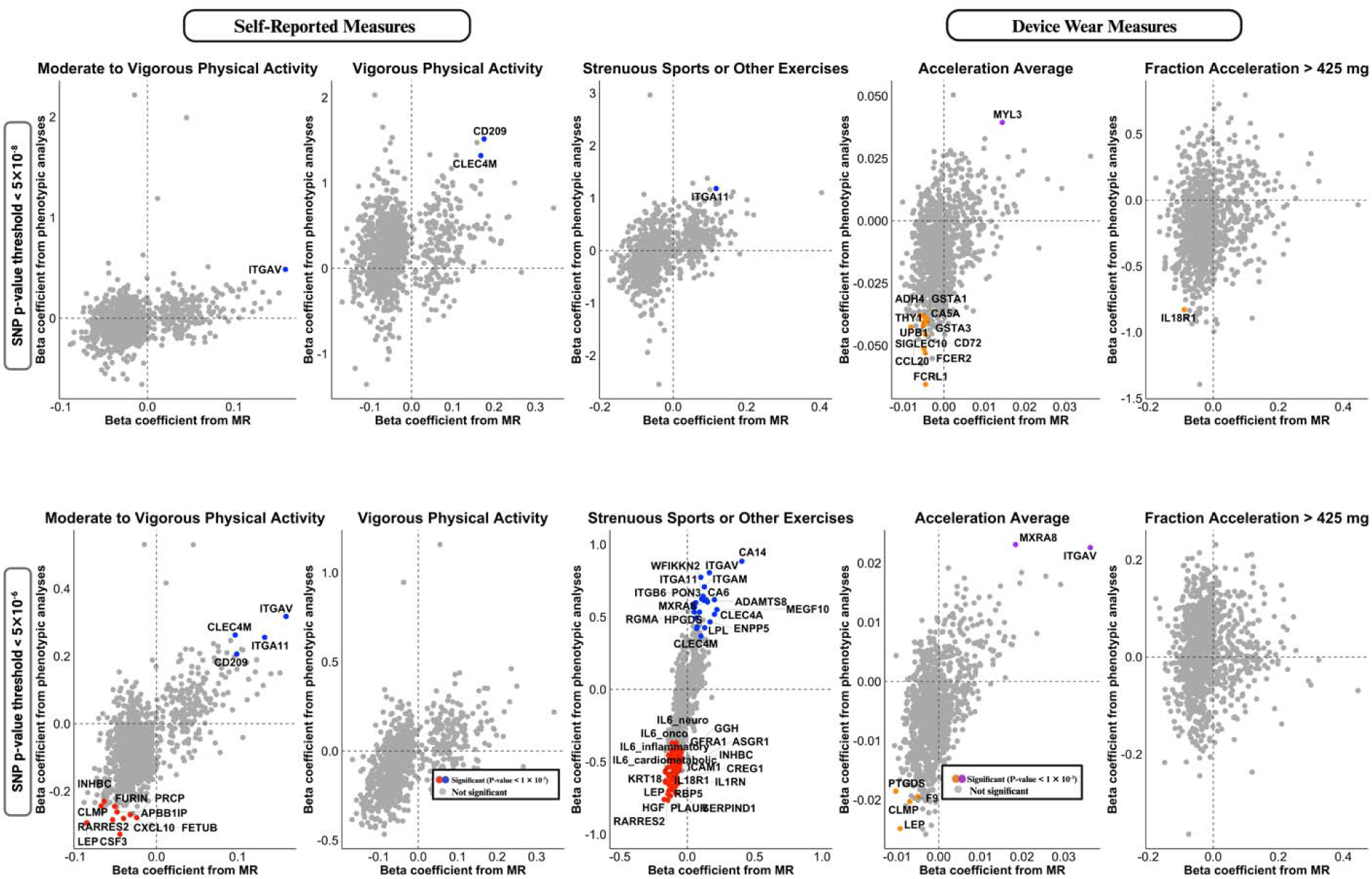
Comparison of observational and Mendelian Randomization effect estimates for the associations between PA exposures and protein levels. The figures demonstrate the relationship between beta coefficients derived from conventional observational associations (y-axis) and MR analyses (x-axis) for self-reported PA measures (Moderate to Vigorous Physical Activity, Vigorous Physical Activity, Strenuous Sports or Other Exercises) and device-wear measures (Acceleration Average, Fraction Acceleration > 425 milligravities). Each colored data point corresponds to a protein, with significant associations in both the observational and MR analyses (p < 1 × 10 ^-3^) highlighted. These plots offer a visual comparison to evaluate the consistency in effect estimates across both observational and MR analyses. (Created in part with <u>BioRender.com</u>)

### Mediation and interaction analyses

There were up to 859 incident dementia cases identified among participants with both proteomic and PA data (Supplementary Table S24). Our investigation employing the 4-way decomposition method identified 15 proteins that significantly mediated the relationship between PA and incident dementia (p < 4.9 × 10^-5^). Five proteins were found to mediate the relationship between MVPA and dementia; three proteins mediated the relationship between VPA and dementia, and five proteins mediated the relationship between SSOE and dementia (Table 2; Supplementary Tables S25). We did not identify any protein as a significant mediator of the effect of device-based measures on dementia.

**Table 2.** Mediation analysis results show the proteins that significantly mediated the relationship between PA and all-cause dementia.

| PA measure | Protein | Pure indirect effect | P<sup>1</sup> |
| --- | --- | --- | --- |
| **MVPA** |  |  |  |
|  | GDF15; Growth/differentiation factor 15 | -0.01 [-0.02, 0.01] | 1.69E-07 |
|  | HPGDS; Hematopoietic prostaglandin D synthase | -0.01 [-0.02, -0.01] | 4.81E-07 |
|  | ITGAV; Integrin alpha V | -0.02 [-0.02, -0.01] | 8.38E-07 |
|  | MENT; Protein MENT | 0.01 [-0.02, -0.01] | 3.06E-06 |
|  | ITGAM; Integrin alpha M | -0.01 [-0.02, -0.01] | 7.31E-06 |
| **VPA** |  |  |  |
|  | ITGAV; Integrin alpha V | -0.06 [-0.08, -0.03] | 2.62E-06 |
|  | HPGDS; Hematopoietic prostaglandin D synthase | -0.04 [-0.06, -0.02] | 3.28E-06 |
|  | GDF15; Growth/differentiation factor 15 | -0.04 [-0.06, -0.02] | 1.18E-05 |
| **SSOE** |  |  |  |
|  | GDF15; Growth/differentiation factor 15 | -0.05 [-0.06, -0.03] | 8.38E-08 |
|  | HPGDS; Hematopoietic prostaglandin D synthase | -0.03 [-0.04, -0.03] | 6.52E-07 |
|  | ITGAV; Integrin alpha V | -0.03 [-0.05, -0.02] | 1.13E-05 |
|  | ITGAM; Integrin alpha M | -0.02 [-0.03, -0.01] | 3.27E-05 |
|  | BCAN; Brevican core protein | -0.02 [-0.03, -0.01] | 3.38E-05 |

**Pure indirect effect (PIE):** This represents the estimated indirect effect of the protein on the relationship between PA and dementia based on the 4-way decomposition, with 95% confidence intervals provided in parentheses. **P:** The p-value for the pure indirect effect<sup>1</sup> Presented here are the protein mediators that are statistically significant after correction for multiple testing ( $p < 4.9 \times 10^{-5}$ )

Growth Differentiation Factor 15 (GDF15), Hematopoietic Prostaglandin D Synthase (HPGDS) and Integrin Subunit Alpha V (ITGAV) demonstrated consistent mediating effects across MVPA, VPA, and SSOE on dementia risk. Integrin Subunit Alpha M (ITGAM) mediated the effects of MVPA and SSOE on dementia risk. Glial Fibrillary Acidic Protein (GFAP) showed mediating effects of MVPA and VPA on dementia. Brevican core protein (BCAN) was also shown to mediate the effect of SSOE on dementia.

## DISCUSSION

In this study, we aimed to explore how PA influences the plasma proteomic landscape and to identify proteins that mediate PA’s protective effect on dementia. We conducted thorough analyses across 2,911 proteins to identify consistent findings across different PA measures and study designs. Conventional observational analyses revealed 1,027 proteins significantly associated with at least one of the six PA measures, and 41 proteins significantly associated with all six PA measures. We leveraged bi-directional MR analyses to validate and enhance the robustness of our findings regarding the putative causal effects of PA on protein levels. Pathway enrichment analyses revealed several biological pathways that may be upregulated in response to PA, and mediation analyses revealed specific proteins that may mediate the relationship between PA and dementia.

Our study extends previous findings by replicating several proteins such as FAP, LEP, CD209, CD248, MXRA8, WFIKKN2, ADM, APOA1, and IL6.^18,21,24,25^ We also identified many novel proteins and evaluated the evidence for causality and direction of effect between PA and each protein using a genetically-informed study design. The identified proteins fall into several broad functional categories, including cardiovascular and vascular regulation (e.g., ADM, ITGB5), lipid metabolism (e.g., APOA1, LPL), inflammatory and metabolic regulation (e.g., DPP4, GDF15, HPGDS), and tissue remodeling and fibrosis (e.g., FAP), highlighting the diverse physiological mechanisms through which PA influences critical cellular functions.^47^

Across conventional observational, MR, mediation, and pathway enrichment analyses, we consistently identified the integrin family of proteins (e.g. ITGAV, ITGAM, and ITGA11) as increasing in response to PA, decreasing in response to SB, and mediating the effect of PA on all-cause dementia. Integrins are transmembrane receptors involved in cell adhesion and signaling and have been shown to significantly influence the extracellular matrix (ECM), critical for maintaining tissue integrity and facilitating cellular interactions.^48^ Integrins such as ITGAV, ITGAM, and ITGA11 were consistently associated across multiple PA measures, underscoring the role of PA in ECM remodeling and cell-extracellular matrix adhesion. Another protein that we consistently identified was MXRA8, which is also involved in ECM processes.^49^ MXRA8 may interact with integrins such as ITGAV, playing a role in the maturation and maintenance of the blood-brain barrier.^49^ Integrins could also explain the potential roles of PA in reducing the risk of all-cause dementia and Alzheimer’s disease via facilitating cell-extracellular matrix adhesion functions that impact neuronal signaling pathways, synaptic plasticity, neuroinflammation, and blood-brain-barrier integrity.^50^ Additionally, integrin-mediated pathways, such as those involving ITGA2β1 and ITGAVβ1, have been found to reduce amyloid-beta toxicity, further linking the positive effects of PA with brain health and reduced neurodegenerative risk.^51^ Finally, integrins are strongly linked to irisin, a previously identified exerkine.^52,53^ Irisin facilitates neurogenesis and amyloid-beta degradation through integrin receptors, such as ITGAVβ5, on astrocytes and may also be involved with BDNF activation via integrin receptor complexes.^52,53^

We also found that proteins such as IL-6, HPGDS, and GDF15 were consistently associated with PA and SB, implicating the modifications of inflammatory, metabolic, and vascular processes.^54,55^ Previous studies have indicated that IL-6 and GDF15 levels may transiently rise following acute PA, but tend to decrease with sustained long-term PA, suggesting that our measurements may capture these lasting benefits effectively.^56–60^ HPGDS was previously found to have a role in dementia via inflammatory pathways.^55^ Notably, HPGDS and GDF15 also emerged as significant mediators between PA and the risk of all-cause dementia, suggesting that PA protects against dementia by reducing inflammation. The findings reinforce the broader evidence supporting the involvement of these proteins in neurodegenerative pathways.^61^ The increase of HPGDS via PA could be related to prostaglandin-mediated inflammatory processes implicated in neurodegeneration.^55^ Elevated GDF15 has been previously associated with increased risks of all-cause dementia, vascular dementia, and Alzheimer’s disease, particularly in the context of cerebrovascular damage.^54,61,62^ The reduction in GDF15 that we observed with long-term PA suggests an anti-inflammatory compensatory response in the brain, which may minimize damage and promote recovery.^61,62^ These results underscore the need to explore further associations between PA and these inflammatory markers and their role in vascular health, reducing neuroinflammation and promoting neuronal resilience.

Other proteins that were consistently associated with PA included LPL, DPP4, FAP, LEP, and IGFBP1, which may play roles in cardiometabolic diseases and cancer. LPL, a protein that breaks down triglycerides into free fatty acids for energy, is increased with PA, which might help improve lipid profiles.^63–65^ Another protein, FAP, which is primarily involved in tissue remodeling and fibrosis, was found to increase with PA. In previous research, elevated FAP expression after exercise correlated with improved cardiovascular metrics, such as VO_2_ max, suggesting FAP’s possible role in musculoskeletal tissue repair and cardiovascular health.^25,66–69^ Moreover, we also identified LEP, a well-known metabolic hormone that regulates energy balance and satiety.^70^ It consistently decreased with PA after controlling for percent body fat. This supports the previously reported role of PA in regulating appetite, possibly via LEP.^71,72^ IGFBP1, which increases with PA, regulates insulin-like growth factors essential for glucose metabolism and cell growth.^73^ Elevated IGFBP1 levels aid in better insulin regulation and metabolic stability, which may be crucial for preventing type-2 diabetes and supporting tissue repair.^73,74^ Finally, we found that EGFR, another protein known for its role in cell growth and differentiation, increases with PA, which may help reduce cancer risk by preventing aberrant cellular proliferation.^75^ Although PA has been studied as a protective factor for cancer, the relationship between PA and EGFR is not well understood, warranting further study.^76^

Our study is notable because it is the largest and most comprehensive study of PA and plasma protein levels. Our approach included various self-reported and device-based PA measures, which allowed us to identify proteins with the most consistent associations across these measurement types, and proteins that exhibited associations with only a subset of measures. Employing a genetically-informed study design enabled us to examine the existence and direction of causal links between proteins and PA. Thus, specific proteins with consistent magnitudes and directions of association across study designs and PA measures could be identified. To assess the mediation of the PA-dementia relationship through specific proteins, we employed the 4-way decomposition method to disentangle the distinct contributions of PA’s direct, indirect, and interactive effects on dementia through a given protein. This approach facilitated the identification of specific pathways and mechanisms that could underlie the protective effect of PA.

However, analyses of device-based measures in our conventional observational design may be vulnerable to temporal ambiguity and potential reverse causation since there is a 4–8-year time gap between baseline blood sample collection and the device-based measure. Our bi-directional MR analyses could provide some clarity as to any potential reverse causation. Mediation analyses might be prone to residual confounding and temporal ambiguity for the device-based measures, as mentioned above. Moreover, in mediation and interaction analyses, the total effect is only represented by the exposure and mediator, making strong assumptions about unaccounted confounding and the independence of protein mediation. The study population primarily consists of middle-aged individuals of European descent, which limits the generalizability of the findings to other ancestries and age groups. Additionally, the MR approach relies on several assumptions, some of which are challenging to test. The gene-environment equivalence assumption posits that the downstream effects of genetic variants on a given exposure are like those of the environmental exposure itself, which may not always hold. Pleiotropy, as indicated in some cases by the colocalization analyses, is another assumption that may not hold.

We identified several proteins influenced by PA that span functions such as cell adhesion, inflammation, and cardiometabolic and neuroprotective roles. By integrating proteomic data with detailed epidemiological information and various study designs, our study provides novel insights into how PA affects health at the molecular level, potentially guiding precision public health interventions. These proteins reveal biological mechanisms that warrant closer investigation, especially in their role as molecular transducers of PA. The findings could inform strategies for tracking and enhancing PA behaviors and highlight biological pathways linked to dementia, which may be prioritized for therapeutic intervention.

## Data Availability

SUPPLEMENTARY MATERIAL
Supplementary tables can be found in our GitHub repository: https://github.com/klimentidis-lab/ProteomicsofPhysicalActivity2024.git
DATA AND CODE AVAILABILITY
Individual-level data from the UK Biobank can be requested at https://www.ukbiobank.ac.uk/enable-your-research/apply-for-access. Summary statistics for all GWAS used in this study are publicly available from the GWAS catalog (64) and UKB-PPP at https://doi.org/10.7303/syn51364943. Our supplementary tables, analysis codes, and results including interactive visualizations of results, are available in a GitHub repository: https://github.com/klimentidis-lab/ProteomicsofPhysicalActivity2024.git

https://github.com/klimentidis-lab/ProteomicsofPhysicalActivity2024.git

## SUPPLEMENTARY MATERIAL

Supplementary tables can be found in our GitHub repository: https://github.com/klimentidis-lab/ProteomicsofPhysicalActivity2024.git

## DATA AND CODE AVAILABILITY

Individual-level data from the UK Biobank can be requested at https://www.ukbiobank.ac.uk/enable-your-research/apply-for-access. Summary statistics for all GWAS used in this study are publicly available from the GWAS catalog (^64^) and UKB-PPP at https://doi.org/10.7303/syn51364943. Our supplementary tables, analysis codes, and results including interactive visualizations of results, are available in a GitHub repository: https://github.com/klimentidis-lab/ProteomicsofPhysicalActivity2024.git

## ACKNOWLEDGMENTS

This research has been conducted using the UK Biobank Resource under Application Number 21259. It uses data provided by patients and collected by the NHS as part of their care and support. The authors thank the UK Biobank’s organizers and participants.

## FUNDING

The authors would also like to acknowledge funding from NIH R01AG072445.

## CONFLICT OF INTEREST

The authors have no conflict of interest to report.

## Notes

### Competing Interest Statement

The authors have declared no competing interest.

### Author Declarations

Ethics committee/IRB of the North West Multi-centre Research Ethics Committee (MREC) waived ethical approval for this work under UK Biobank's Research Tissue Bank (RTB) approval.

### Summary of Updates

We corrected the misspecified author names, addressed grammatical issues in the text, and adjusted the layout of the figures and tables.

